# Muscle and Lipid Suppression: *MUSLIS* MRI, a novel approach to musculoskeletal MRI

**DOI:** 10.64898/2026.10.11.26364852

**Authors:** Giulio Gambarota, Jérémy Beaumont, Mark Tanner

## Abstract

**Purpose:** to introduce a novel magnetic resonance imaging (MRI) technique, **MUSLIS** MRI (**mus**cle and **li**pid **s**uppression), that generates two co-registered 3D sets of images in a single acquisition: one set with lipid signal suppression (MUSLIS1) and a second set with muscle signal suppression (MUSLIS2).

**Materials and Methods:** Numerical simulations based on Bloch equations were performed to optimize the sequence parameters; the MP2RAGE sequence was employed for the implementation of MUSLIS imaging. MRI experiments were performed *in vivo* at 3 T. Quantitative measurements of signal contrast were carried out on MUSLIS1 and MUSLIS2. Various combination images and T1 parametric maps of the thighs were generated from the two co-registered 3D sets.

**Results and Discussion:** Optimal sequence parameters to achieve lipid suppression in one dataset (MUSLIS1) and muscle suppression in the other dataset (MUSLIS2) were determined by numerical simulations. MUSLIS imaging was validated *in vivo* at 3 T: MUSLIS1 images and MUSLIS2 images displayed excellent lipid and muscle suppression, respectively. The values of the signal contrast between subcutaneous lipids and muscle, as well as between bone marrow lipids and muscle reached values close to 0.8. Thus, MUSLIS would allow for tissue characterization of lesion/masses and direct visualization of the adipose tissue. Combination images, generated from these two intrinsically co-registered sets of images (MUSLIS1 and MUSLIS2), displayed different contrasts and improved signal characteristics, such as mitigation of signal inhomogeneities. The development of MUSLIS imaging for musculoskeletal imaging followed the concept introduced with the FLAWS, an MRI sequence designed for brain MR imaging to generate two 3D sets of images, one with white matter suppression and the other with cerebrospinal fluid suppression. Similarly to FLAWS, which provides T1 maps of the brain, MUSLIS imaging yields T1 parametric maps of the musculoskeletal system. Furthermore, the unique characteristics of the combination images make MUSLIS imaging a potentially attractive technique at field strengths higher than 3 T.

**Conclusion:** The proof-of-concept of a novel approach to musculoskeletal imaging, MUSLIS MRI, is proposed here and demonstrated by numerical simulations and MRI experiments performed *in vivo* at 3 T.

**Graphical Abstract:** 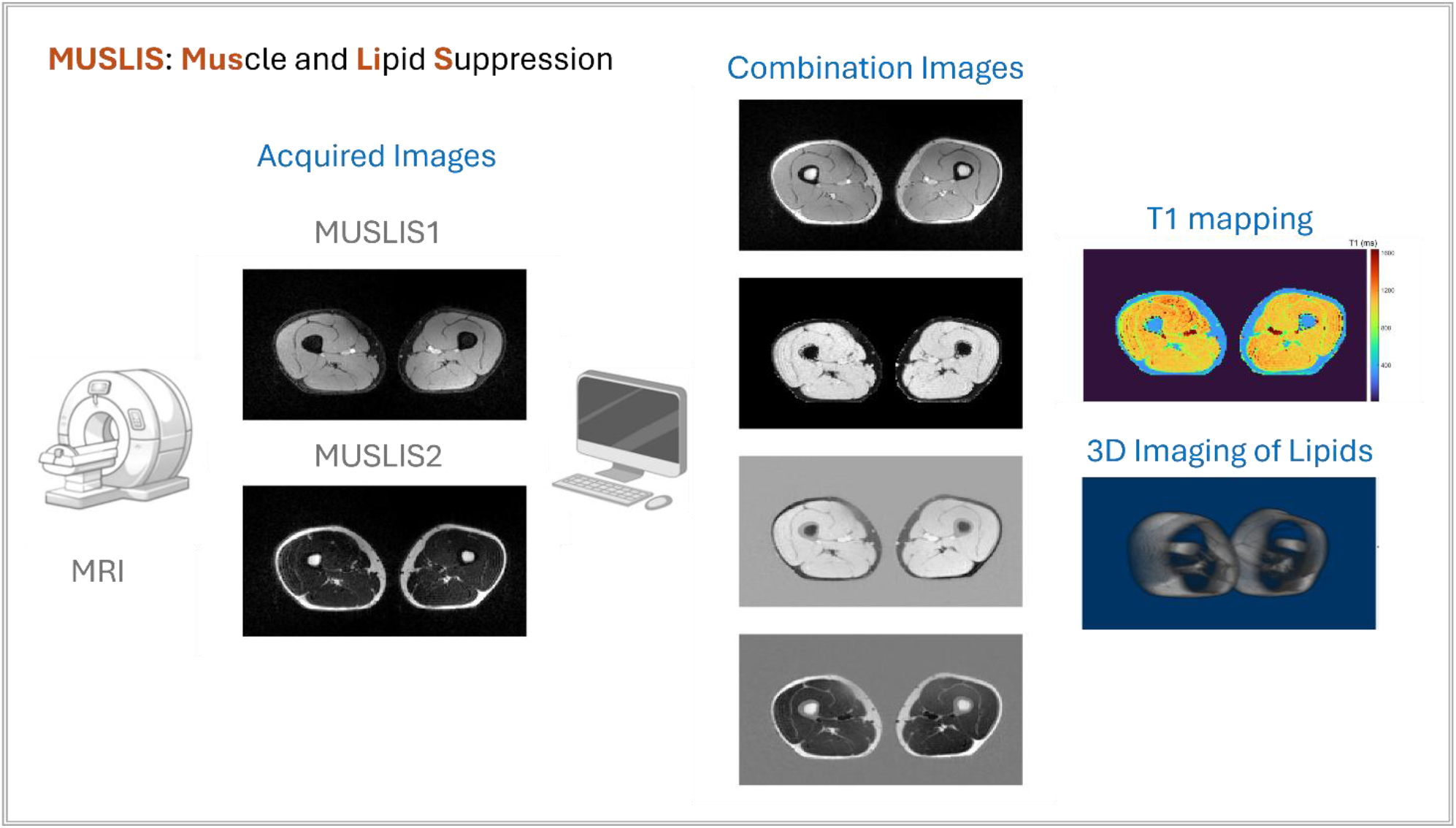

## Introduction

### MRI, an essential technique for MSK studies

Magnetic resonance imaging (MRI) is a fundamental technique for many investigations of the musculoskeletal (MSK) system, both in clinical practice and research (1–5). A standard musculoskeletal MRI protocol consists of the acquisition of different types of images, such as T1-weighted (T1w) and T2-weighted (T2w) images, which yield different contrasts between tissues and thus contribute to the diagnosis of pathological conditions (6–12). Each type of acquisition can serve one or more purposes; for instance, T1w images allow for anatomical delineation of muscles and other musculoskeletal tissues; T2w images highlight tissue regions affected by inflammation processes. Furthermore, it should also be noted that an MSK protocol includes acquisitions of images where the signal of lipid is “suppressed” (13–15), as we will detail in following sections.

### Muscle and adipose tissue

Among the tissues of the MSK system, muscle and adipose tissue have an important role, especially in non-traumatic MSK pathologies. Adipose tissue (i.e., lipids, or fat) is present in various compartments of the musculoskeletal system, including: bone marrow fat, subcutaneous fat, fat depots between muscle fibers or muscle groups, and fat droplets stored within myocytes (16–26). Furthermore, adipose tissue might also be present in both tumoral and non-tumoral lesions (27–37).

### MRI and lipid suppression

A typical MSK MRI protocol includes the acquisition of at least one imaging dataset where lipid signal is suppressed (*lipid suppression* is also referred to as *fat suppression*). In general, the signal of lipids might i) hinder visualization of important pathophysiological processes - as inflammation, for instance- and ii) impede lesion characterization, that is, hamper the tissue content evaluation of lesions/suspicious masses. Thus, the acquisition of images where the lipid signal is “suppressed” is an essential part of an MSK MRI protocol.

### Lipid suppression techniques

There are different strategies to suppress the lipid signal in MRI: all strategies exploit the differences in magnetic properties between the hydrogen (^1^H) nuclei of water (also referred to as water protons) and those of lipids. One of the main differences is the value of the T1 relaxation time: the T1 of lipids is much shorter than that of water in most tissues. Historically, the first approach was indeed the ‘Short Tau Inversion Recovery’ sequence (STIR, where tau is a time delay, also referred nowadays as TI = inversion time), which exploits this difference in T1 relaxation time (38,39). The STIR sequence starts with an inversion pulse that generates a negative longitudinal magnetization of both water and lipids; the magnetization recovery (i.e., the relaxation to the equilibrium state) of lipids is faster than that of water, since the T1 of lipids is shorter. Following the inversion pulse, the signal is acquired after a short time delay (= short TI), when the lipid longitudinal magnetization goes through the zero value. Other lipid suppression techniques exploit the differences in resonance frequency between the hydrogen nuclei of water and lipids (i.e., the water-fat chemical shift), by using frequency selective radiofrequency pulses (40–42); the most recent techniques (‘hybrid’ techniques) combine the STIR approach with frequency selective radiofrequency pulses (13,14,43).

### Inversion recovery techniques

The STIR sequence belongs to the class of MRI sequences named inversion recovery (IR) techniques. Since the beginning of the MRI development in the 1980s, scientists have exploited the benefits of this type of sequences for different applications (38,39). In brain imaging, it is of interest to suppress the signal of cerebrospinal fluid (CSF) in order to better identify gray matter (GM) and white matter (WM) structures and measure the volume/shape of these structures, for instance. One major difference in magnetic properties among water protons of GM, WM and CSF is the very long T1 of CSF. CSF suppression is implemented in i) the FLAIR sequence, which typically share with the STIR sequence the fast spin echo acquisition method, and in ii) the MPRAGE sequence, which has a spoiled gradient acquisition module instead (FLAIR: Fluid-Attenuated Inversion Recovery; MPRAGE: Magnetization-Prepared Rapid Acquisition Gradient Echoes) (44–47).

### From MPRAGE to FLAWS, from STIR to MUSLIS

In recent years, an ‘expanded’ version of the MPRAGE (the MP2RAGE, Magnetization-Prepared 2 Rapid Acquisition Gradient Echoes) was developed to improve brain image quality at high magnetic fields (7 T) and to provide T1 maps of the brain (48). The MP2RAGE allows for the simultaneous acquisition of two datasets of images that, when combined, yield an image dataset with reduced bias field effects (49). Following the introduction of the MP2RAGE, a novel brain imaging approach was proposed, the FLAWS (FLAWS: FLuid And White matter Suppression, Table 1). In FLAWS imaging, the choice of sequence parameters (in particular, the inversion times) yields i) FLAWS1, characterized by WM suppression and ii) FLAWS2, with the CSF suppression -as a standard MPRAGE (50). FLAWS allows for better delineation of subcortical brain structures, such as the external and internal globus pallidus, which is important in a several applications such as deep brain stimulation (DBS). Similarly to the evolution from MPRAGE to the FLAWS (Table 1) we sought to expand the STIR (lipid suppression) approach to a novel sequence, **MUSLIS** (**mus**cle and **li**pid **s**uppression), that yields i) an image dataset with lipid suppression (**MUSLIS1**) and ii) another image dataset with muscle suppression (**MUSLIS2**). MUSLIS1 would allow for tissue characterization of lesion/masses and for direct 3D imaging of the adipose tissue. Furthermore, the simultaneous acquisition of two co-registered datasets of images allows for generating i) new datasets with different contrasts and improved image quality and ii) T1 parametric maps, as in FLAWS imaging.

**Table 1.** From MPRAGE to FLAWS, from STIR to MUSLIS: ‘standard’ vs ‘newly proposed’ MR inversion recovery techniques for brain imaging and MSK imaging. **\***It should be pointed out the difference between the acquisition method of STIR (fast spin echo) vs MUSLIS (rapid gradient echo).

| Brain imaging | MPRAGE | FLAWS |
| --- | --- | --- |
| Signal Suppression | CSF | CSF (FLAWS2) |
|  |  | White Matter (FLAWS1) |
| MSK imaging | STIR | MUSLIS* |
| Signal Suppression | Lipids | Lipids (MUSLIS1) |
|  |  | Muscle (MUSLIS2) |

## Materials and Methods

### MUSLIS imaging, the implementation: simulations and *in vivo* imaging

Similarly to the FLAWS imaging development (50), the implementation of MUSLIS imaging was carried out using the MP2RAGE sequence (48), a vendor-provided sequence available on Siemens MR clinical systems. Bloch simulations of the MUSLIS signal were performed with the goal of identifying the optimal sequence parameters to achieve lipid suppression in one dataset and muscle suppression in the other dataset. Briefly: the MP2RAGE consists of an adiabatic inversion pulse, followed by two gradient-echo (GRE) readout modules to generate two 3D sets of images: one at a first inversion time (*TI*_1_) and the other at a second inversion time (*TI*_2_). Thus, the signal of MUSLIS is a function of the following sequence parameters:

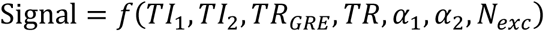

where: *TI*_1_ and *TI*_2_ are the inversion times, *TR* the MP2RAGE repetition time (the time interval between two consecutive inversion pulses), *TR_GRE_* the repetition time of the GRE modules, *α*_1_, *α*_2_ the flip angles of the GRE modules, *N_exc_* the number of excitations per GRE module. Combinations of all parameters were investigated to obtain a parameter set that yields lipid suppression in one 3D dataset and muscle suppression in the other 3D dataset. The literature values of T1 of lipid and muscle at 3 T were used in the simulations (51). MRI experiments were performed *in vivo* at 3 T (Magnetom Verio, Siemens Healthineers, Erlangen, Germany) on one volunteer, with a 24-element spine matrix coil and a flex coil, which was positioned over both legs. The MUSLIS sequence parameters were: TR = 3000 ms, TI1 = 250 ms, TI2 = 700 ms, flip angles: 5° and 5°, acquisition matrix = 114x192, number of slices = 52, bandwidth = 400 Hz/px, FOV = 249*420 mm^2^; voxel size = 2.2x2.2x2.2 mm^3^. The field of view included both legs; the imaging volume was placed approximately at the midpoint of the subject’s thighs. The output of the MUSLIS sequence consisted of two sets of images: the 3D volume acquired at TI1 (MUSLIS1) and that acquired at TI2 (MUSLIS2). The experiments were conducted according to the procedures approved by the Institutional Review Board.

### MUSLIS imaging: data analysis

Data analysis was performed with the software Fiji (52). Regions of interest (ROIs) were drawn in muscle, bone marrow and subcutaneous lipids. The contrast (CN) between tissues was calculated according to standard formula *CN_a_*_/*b*_ = |*S_a_* − *S_b_*|/(*S_a_* + *S_b_*), where S is the signal intensity of each tissue.

### MUSLIS imaging: combination images and T1 mapping

Given the fact that MUSLIS1 and MUSLIS2 are *co-registered* 3D datasets, it is possible to generate new 3D datasets with a novel contrast by image combination (50,53–55). MUSLIS-Max is an example of combination image: it is the 3D set obtained by taking, for each pixel, the maximum value between MUSLIS1 and MUSLIS2. In the result section, MUSLIS- Max and various combination images displaying different contrast between tissues are proposed and illustrated. Furthermore, similarly to FLAWS imaging in brain, T1 mapping of the subject’s thighs was performed by applying a previously developed T1 mapping algorithm (48,53).

## Results

### Simulation results and *in vivo* images

Simulations based on Bloch equations were performed for various combinations of imaging parameters. In Figure 1, the results (simulations, upper panel and *in vivo* MR images, lower panel) for a chosen set of parameters are illustrated. In the simulation panel, the yellow bar indicates the value of T1 of lipids (adipose tissue), whereas the green bar indicates the muscle tissue T1; the blue (orange) line illustrates the signal of MUSLIS1 (MUSLIS2) as a function of the T1 of the tissues. The signal characteristics of the acquired *in vivo* MR images closely follow those of the simulated signal. Good lipid suppression and good muscle suppression was observed in MUSLIS1 and MUSLIS2, respectively (see also Figures S1, S2, S3, S4 in the Supplementary Materials).

**Figure 1.**
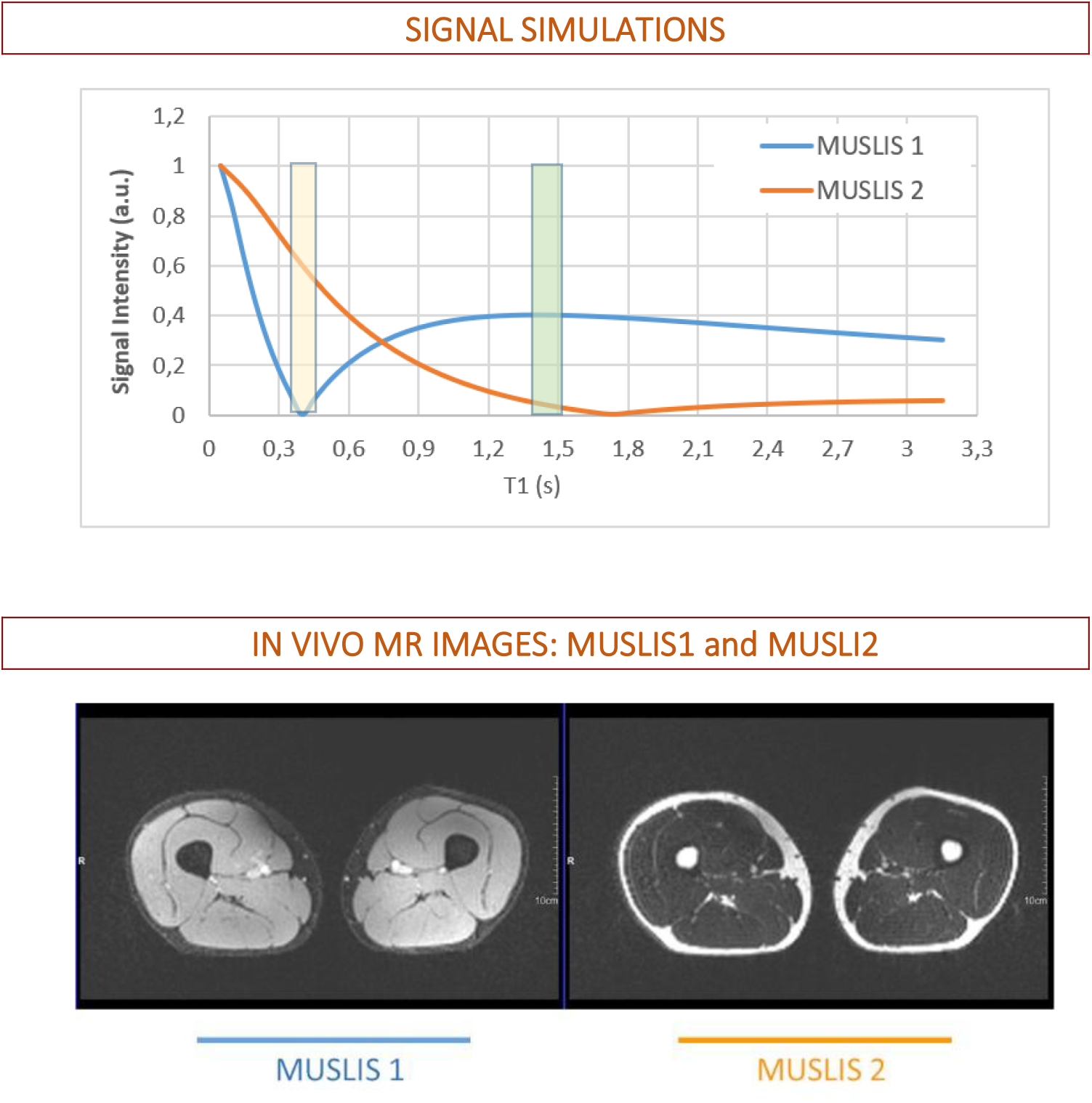
Signal simulations and in vivo MR images of the MUSLIS sequence. **Upper panel**: Simulations of the signal intensity of MUSLIS1 and MUSLIS2 as a function of the T1 relaxation time of tissues. The value of T1 of lipids and muscle (at 3 T: ∼ 0.4 s and 1.4 s, respectively) is highlighted by the yellow box (lipids) and green box (muscle). In MUSLIS1, the signal of lipids is close to zero and the signal of muscle is a relatively high. In MUSLIS2 (the orange line) the signal of muscle (T1 value in the green box) is close to zero) and the signal of lipids is a relatively high. **Lower panel**: In vivo MR images yield similar signal characteristics as the simulations, with good lipid signal suppression in MUSLIS1 (left) and good muscle signal suppression in MUSLIS2 (right).

### MUSLIS imaging and the advantages of co-registered images: the MUSLIS-Max combination image

The first combination image that comes naturally to mind is the one obtained by taking voxel-wise the maximum value between MUSLIS1 and MUSLIS2. The simulations based on Bloch equations and the combination image MUSLIS-Max are shown in Figure 2. MUSLIS-Max results in an image with a high signal intensity of lipids and muscle, as well as a good signal contrast between the two tissues. Overall, MUSLIS-Max has characteristics similar to those of a typical T1w image.

**Figure 2.**
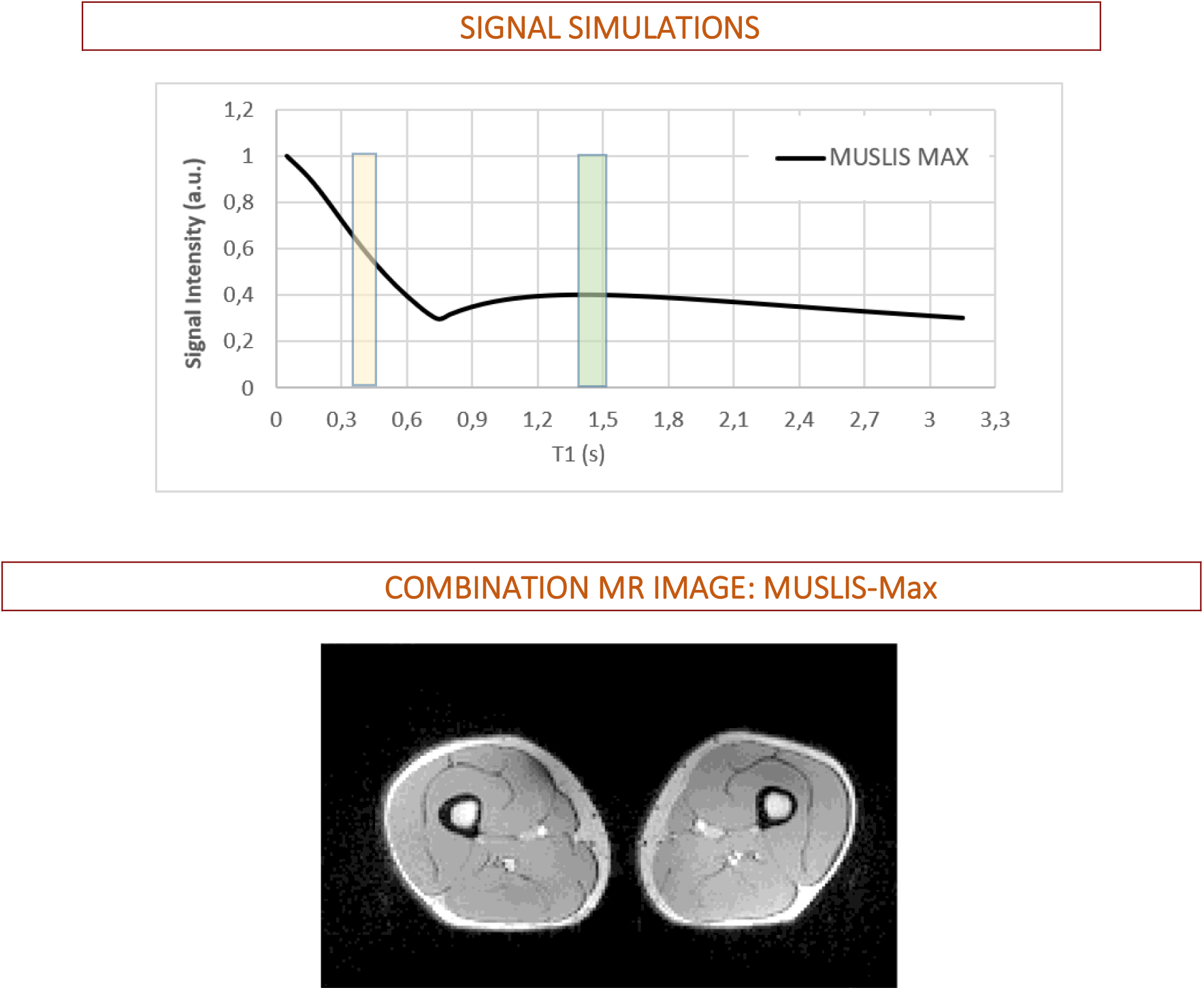
MUSLIS-Max simulations (**upper panel**) and the corresponding MR image (**lower panel**). MUSLI- Max is obtained by voxel-wise combination of the maximum value between MUSLIS1 and MUSLIS2. Both the muscle and lipids are hyperintense in MUSLIS-Max.

### Signal contrast in MUSLIS1, MUSLIS2 and MUSLIS-Max

Quantitative assessment of the signal contrast lipids-muscle was performed and the selected ROIS are indicated in Figures 3, 4, and 5. The signal of lipids was measured in two separate (subcutaneous and bone marrow) compartments. More details and additional measurements of signal contrast are shown in Figures S5, S6, S7 in the Supplementary Materials.

**Figure 3.**
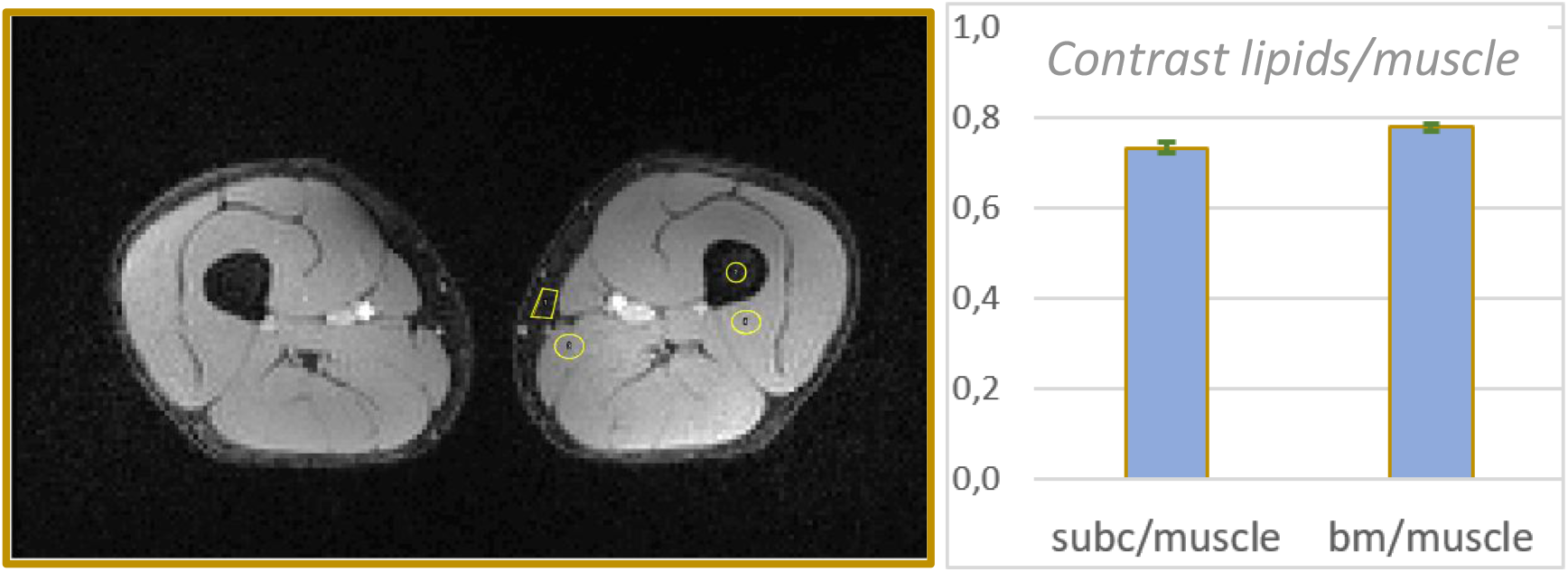
Signal contrast in MUSLIS1. **Left panel**: an example of a MUSLIS1 image and the regions-of-interest (ROIs) located in the subcutaneous (subc) and bone marrow (bm) compartments and adjacent muscles. **Right panel**: the values of the contrast lipids/muscle for the subcutaneous, bone marrow compartment and muscle. The contrast reaches values close to 0.8. Contrast values are averaged over nine contiguous slices; the standard deviation is very small and thus barely visible on the graph.

**Figure 4.**
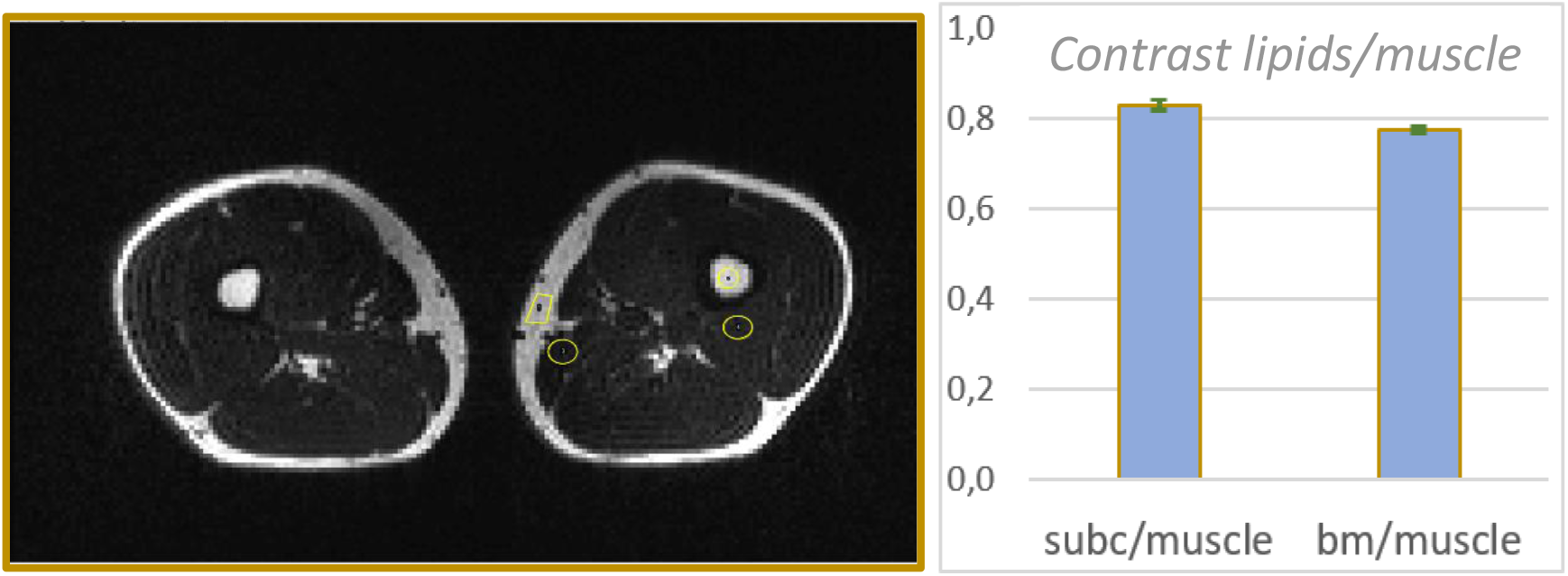
Signal contrast in MUSLIS2. **Left panel**: an example of a MUSLIS2 image and the same regions-of-interest (ROIs) as in MUSLIS1. **Right panel**: the values of the contrast lipids/muscle in MUSLIS2, over nine contiguous slices, are very similar to those of MUSLIS1 (subc = subcutaneous; bm = bone marrow).

### MUSLIS imaging: combination images other than MUSLIS-Max

Examples of signal intensity profiles in MUSLIS1 (acquired image) vs MUSLIS1n (combination image, ‘n’ for ‘normalized’) are shown in Figures 6 and 7. The signal of MUSLIS1n is obtained by the formula *S*_1*n*_ = *S*_1_/(*S*_1_ + *S*_2_), where *S*_1_ (*S*_2_) is the signal intensity of MUSLIS1 (MUSLIS2), with the values of pixels in air set to zero.

**Figure 5.**
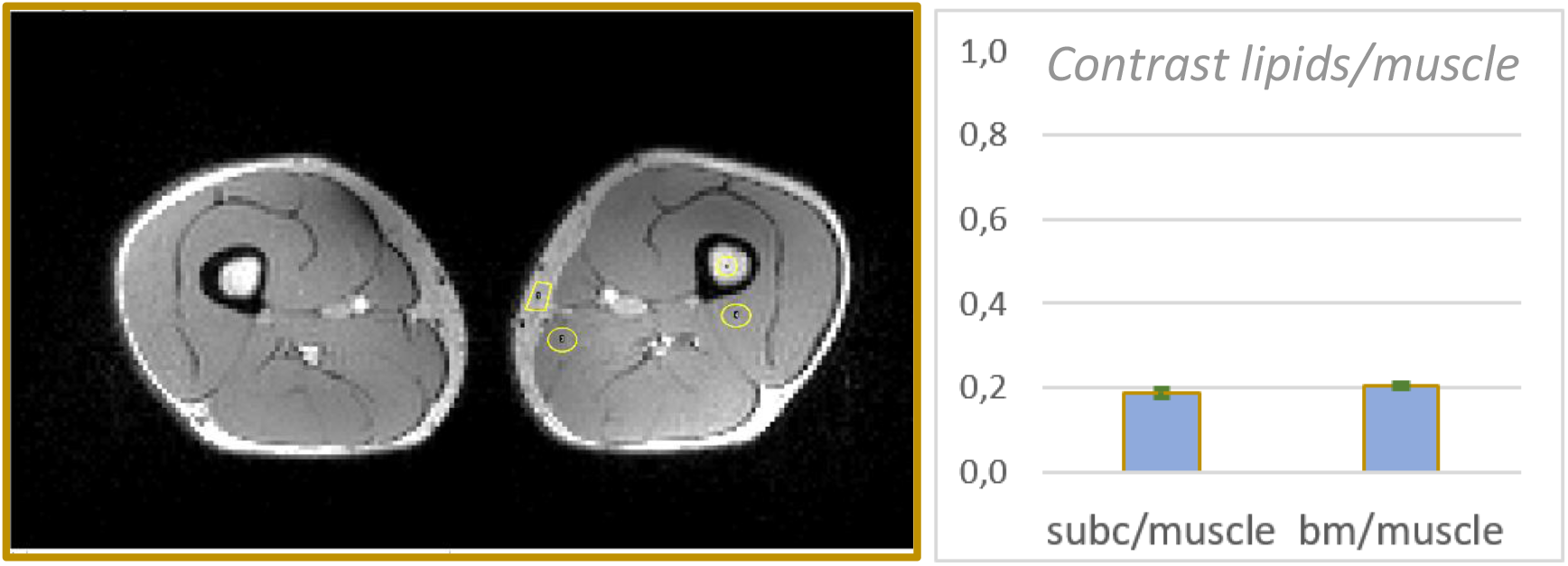
Signal contrast in the combination image MUSLIS-Max. **Left panel**: an example of a MUSLIS-Max image and the same regions-of-interest (ROIs) as in Figure 3 and 4. **Right panel**: the values of the contrast lipids/muscle for the subcutaneous (subc), bone marrow (bm) compartment and muscle.

**Figure 6.**
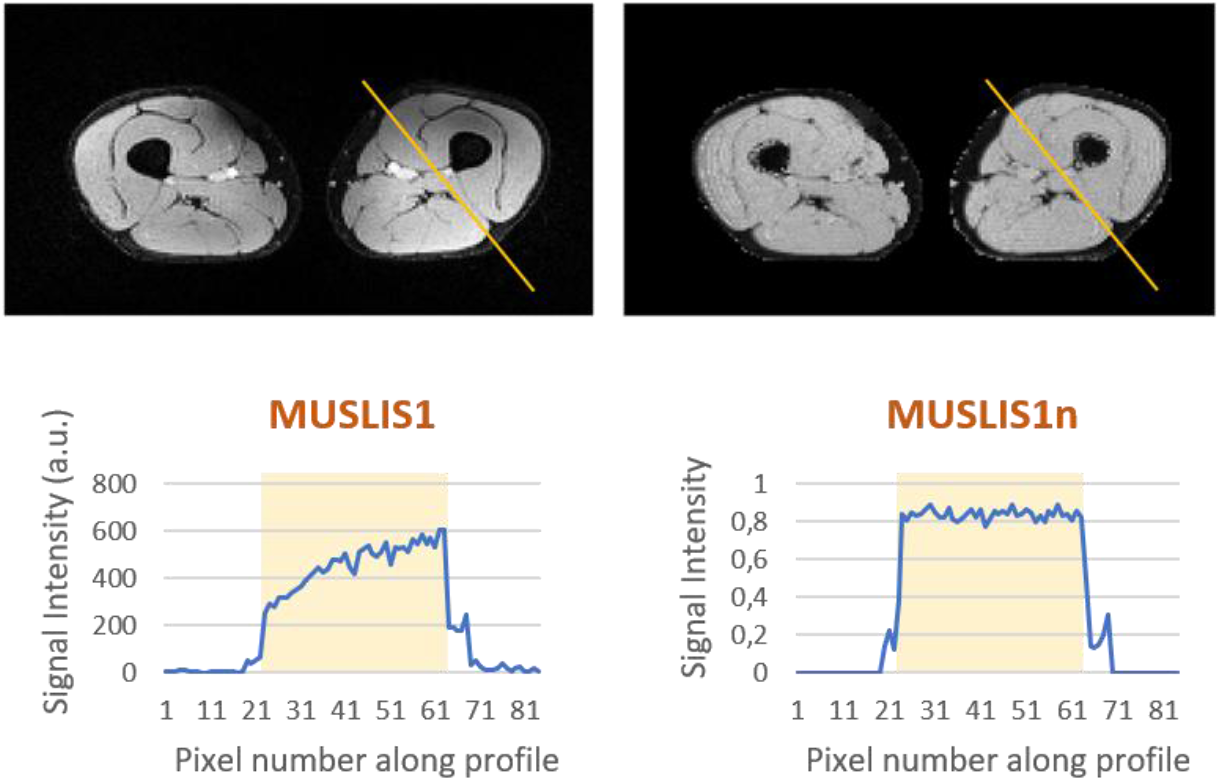
The acquired image MUSLIS1 and the combination image MUSLIS1n (**upper panel**). MUSLIS1n is obtained by dividing the signal intensity of MUSLIS1 by the sum of the signal intensity of MUSLIS1 and MUSLIS2. The signal intensity -along the profile shown on the images- is plotted for MUSLIS1 and MUSLIS1n (**lower panel**). The profile was chosen to cover mostly muscle tissue, trying to avoid other tissue types (bone or lipids, for instance). The yellow box highlights the intensity along the profile, neglecting the extremities of the profile, which include the subcutaneous lipids and air. A substantial improvement in the signal homogeneity along the profile is noticeable in MUSLIS1n. The improvement was quantified by evaluating the value of: i) the average and ii) the standard deviation of the signal along the profile and calculating the coefficient of variation (100* (standard deviation) / average). In MUSLIS1 the coefficient of variation was 18.4% vs 3.2% in MUSLIS1n.

**Figure 7.**
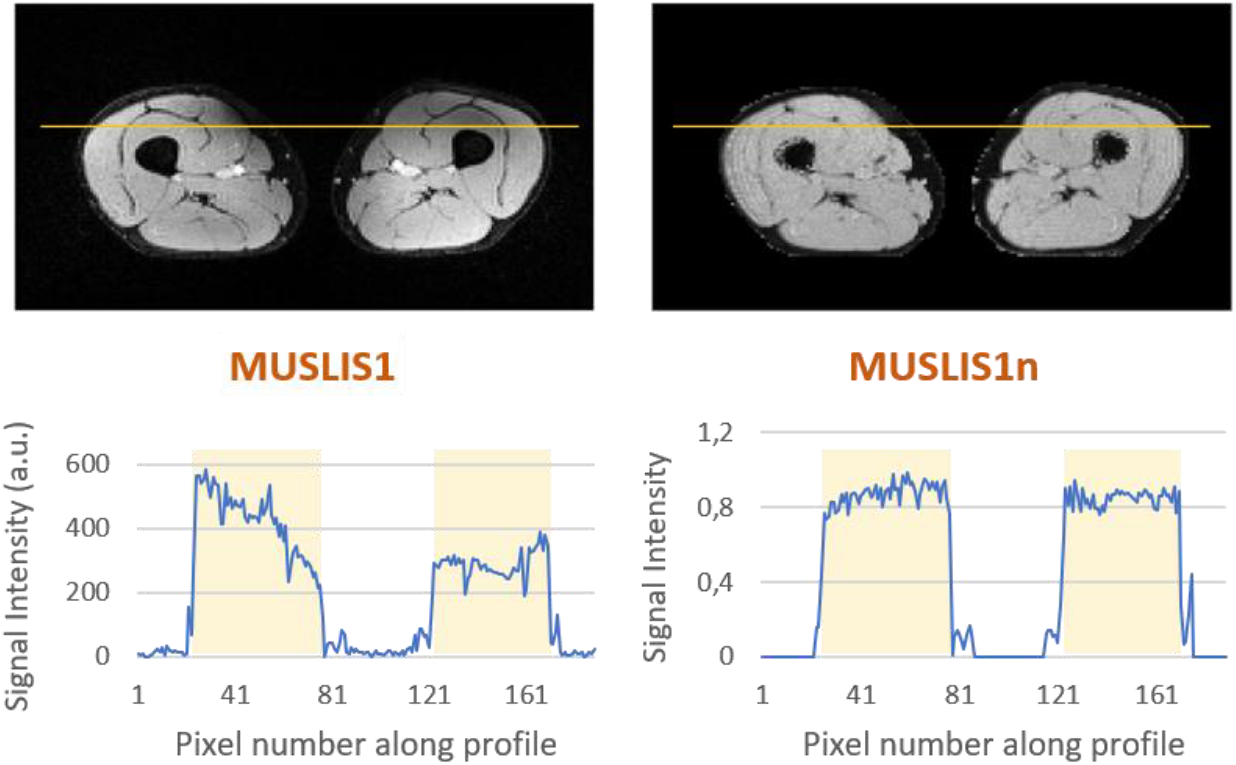
The acquired image MUSLIS1 and the combination image MUSLIS1n (**upper panel**). The signal intensity -along the profile shown on the images- is plotted for MUSLIS1 and MUSLIS1n (**lower panel**). A substantial improvement in the signal homogeneity along the profile is noticeable in MUSLIS1n. See legend of Figure 6 for additional details.

In addition to MUSLIS-Max and MUSLIS1n, other examples of combination images that provide different signal contrast are illustrated in Figure 8: MUSLIS-Diff, with a signal intensity given by the formula *S_diff_* = (*S*_1_ − *S*_2_) and the opposite, MUSLIS-Diff-op: *S_diff_*__*op*_ = (*S*_2_ − *S*_1_) (Figure 8, left column). The normalized version of both is also shown in Figure 8 (right column): to show the effects of the normalization operation on the overall image, the values of pixels in air were not set to zero.

**Figure 8.**
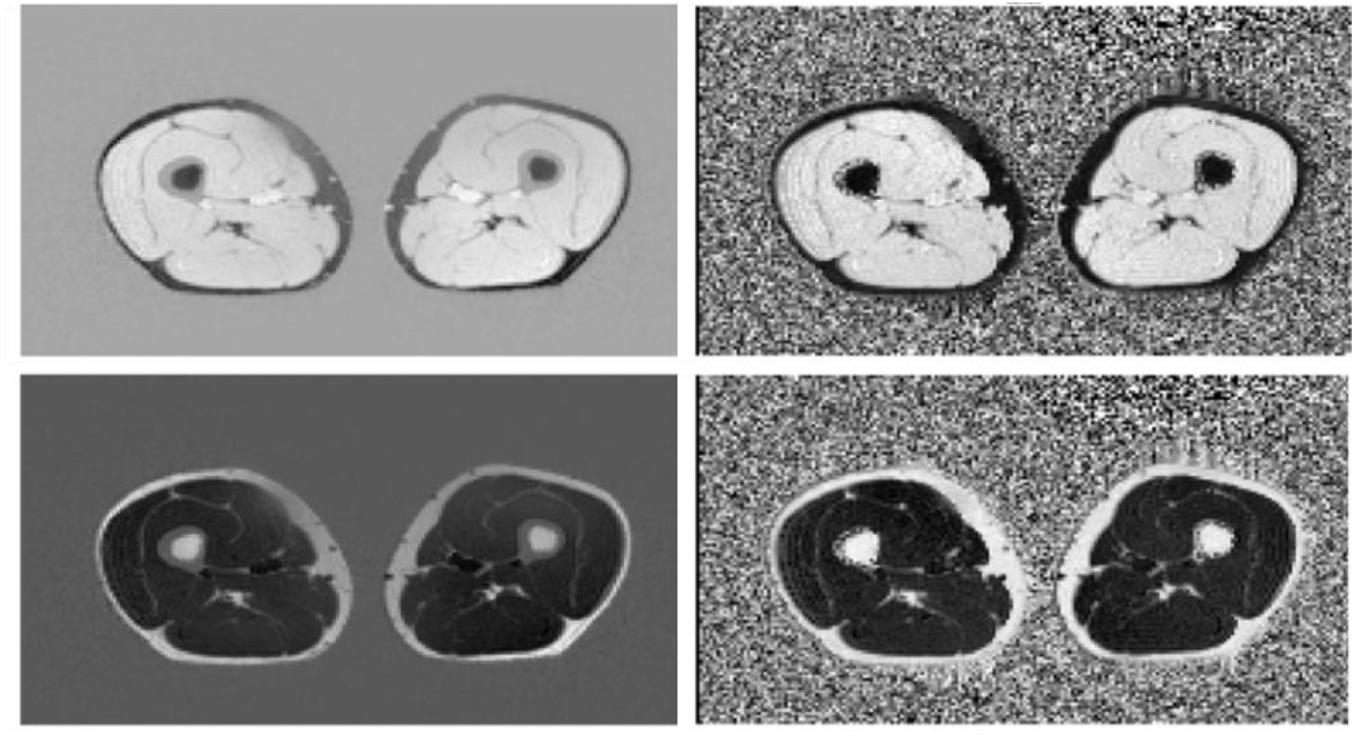
Examples of additional combination images. MUSLIS-Diff and its normalized version is shown in the **upper panel**; MUSLIS-Diff-op and its normalized version is presented in the **lower panel**.

### MUSLIS imaging: additional advantages of 3D imaging

An example of 3D imaging of lipids (MUSLIS2) is shown in Figure 9.

**Figure 9.**
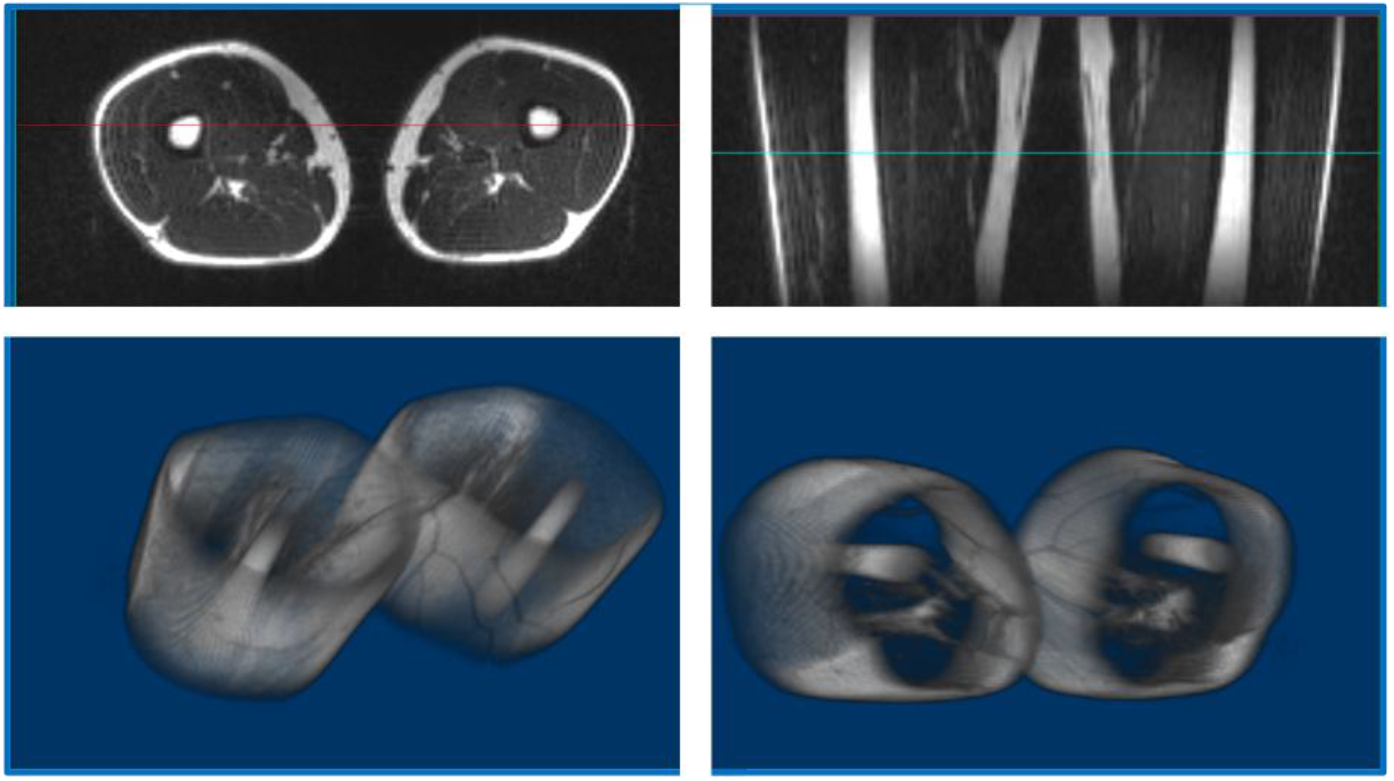
The MUSLIS2 3D image volume, illustrated in an axial and coronal view (**upper panel**) and the 3D visualization of lipids, mostly present in the subcutaneous and bone marrow compartment, in the current volunteer (**lower panel**). In MUSLIS2 images the signal of muscle is suppressed. One of the advantages of the 3D acquisition is the direct visualization of lipids in 3D.

### MUSLIS imaging: T1 parametric maps

MUSLIS imaging employs the MP2RAGE sequence; thus, T1 parametric maps can be obtained from the two acquired images. Figure 10 shows the T1 parametric map of the thighs; for anatomical reference, the corresponding MUSLIS-Max image is also displayed.

**Figure 10.**
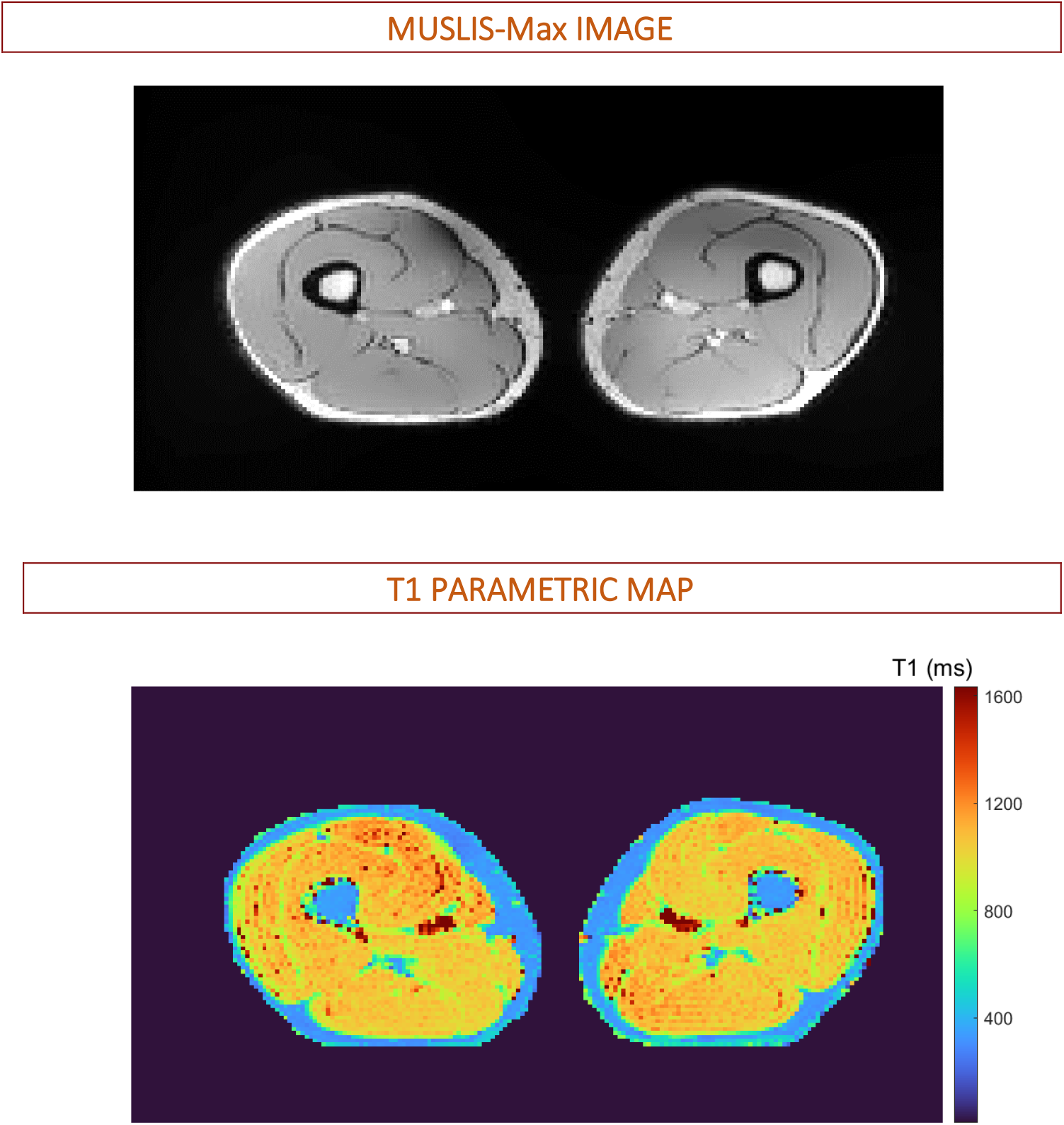
The combination image MUSLIS-Max (**upper panel**), shown for anatomical reference and the corresponding T1 parametric map (**lower panel**). Muscle tissue displays T1 values of approximately 1100 ms. The T1 values of adipose (subcutaneous and bone marrow) tissue are approximately 350 ms.

The analysis of the T1 relaxation times was performed on the slice shown in Figure 10. The T1 of lipids was evaluated in the ROIs located in the subcutaneous and bone marrow compartments, indicated in previous figures (Figure 3, 4 and 5). The T1 was 332±9 ms and 339±20 ms, in subcutaneous tissue, and 344±8 ms and 340±9 ms in bone marrow, for the left and right leg, respectively. With respect to the muscle tissue, the T1 was measured in the ROI that included all muscle tissue in the given slice. The T1 value was 1074±105 ms and 1056±80 ms, for the left and right leg, respectively.

## Discussion

In the current study, we propose a novel approach for MR imaging of the musculoskeletal system, **MUSLIS** (**mus**cle and **li**pid **s**uppression) imaging. In musculoskeletal imaging, the role of lipid suppression is just as important as the ability to acquire images in which only lipids generate a signal (i.e., lipid only images), given the fact that many musculoskeletal pathologies are characterized by fatty infiltration (56–70). With MUSLIS imaging, in a single acquisition, two 3D sets of images are acquired: MUSLIS1 yields images with lipid signal suppression, MUSLIS2 with muscle signal suppression. Thus overall, MUSLIS would allow for lesion identification, tissue characterization of lesion/masses and 3D direct visualization of the adipose tissue.

MUSLIS1 and MUSLIS2 are intrinsically *co-registered* 3D datasets; thus, by image combination it is possible to generate new 3D datasets with multi T1-weighted contrasts and a substantial improvement in signal homogeneity, as shown in Figures 6, 7, 8. These novel images could be useful for segmentation purposes in neuromuscular disorder and metabolic diseases (71–79). In general, it should be noted that recently a number of studies have highlighted the advantages of image combinations (80–84). MUSLIS- Max, for instance, is a standard T1 weighted image. As a result, another advantage of the MUSLIS imaging in MSK could be its potential use following contrast agent (CA) administration. As a matter of fact, in certain cases the MSK protocol might include administration of contrast agent -for instance, to improve specificity in lesion characterization. T1-weighted sequences are employed following CA administration. Therefore, MUSLIS imaging and in particular MUSLIS-Max, which ultimately yields a standard 3D T1-weighted dataset, could be of interest for the acquisition of images post CA administration. Furthermore, the 3D approach of MUSLIS can be an additional advantage for musculoskeletal imaging (85–87). MUSLIS combination images provides also unique advantages: in particular, as in the MP2RAGE and FLAWS approaches, images with reduced bias field effects can be obtained; the results of the current study show major improvements in mitigating signal inhomogeneities at 3 T; this unique characteristic of specific combination images could be of interest at higher fields, such as 7 T.

MUSLIS imaging exploits the difference between the water and lipid T1 relaxation time, to generate separate images of water and lipid (in this context, lipid is also often referred to as fat). A very popular technique that produces fat only images and water only images is based on the difference in the resonance frequency; this is the water-fat separation technique based on Chemical Shift Encoding (CSE), also referred to as Dixon-based technique. As early as 1984, William Thomas Dixon proposed the first example of CSE to generate separate images of fat and water, by acquiring two gradient echo images at different echo times and applying a water/fat decomposition algorithm (88,89). Over the years, many developments of this technique have taken place, with acquisition of multiple echoes (often 6 echoes) and novel algorithms/post processing pipelines to improve robustness of this technique (90–92). Nowadays water-fat separation techniques based on CSE provide i) fat only images, ii) water only images and more importantly iii) fat fraction (often referred to as PDFF, proton density fat fraction) and iv) T2* parametric maps.

The PDFF and T2* are of major interest in liver imaging. The PDFF is useful for diagnosis and follow-up of diseases like steatosis; T2* yields a measure of iron content in liver, which is useful to assess liver iron overload in diseases like hemochromatosis (93–99). MUSLIS imaging provides neither the PDFF nor T2* values; as such, it is of little interest for liver imaging; however, the situation is different in MSK imaging.

With respect to water and fat, a major difference between liver and the MSK system is noteworthy: in liver, fat is present inside the hepatocytes, as lipid droplets (intrahepatocellular lipids); thus the source of fat signal cannot be separated from the source of water signal, they coexist in the same physical space/compartment and the PDFF is essential to evaluate fat accumulation; in the MSK system, the major contribution to the fat signal originates from the adipose tissue, which represents a bulk depot, present in a separate compartment with respect to the muscle tissue. This adipose depot can be measured by MUSLIS. Thus, MUSLIS imaging can play an important role in the diagnosis and follow-up of diseases characterized by fat accumulation in the MSK system. For completeness, it should be also noted that in muscle tissue lipid droplets inside myocytes are present (IMCL, intra-myocellular lipids), but they represent a minor/negligeable contribution compared to the adipose depots; as a matter of fact, IMCL are not visually observable on the MR images - in contrast to the adipose tissue (24–26,100). Furthermore, IMCL can be measured in a somewhat reliable manner only by MR spectroscopic methods, despite methodological efforts in MRI investigations (101). Thus, MUSLIS can be of importance in MSK imaging for assessment of fat in several diseases.

The advantages of MUSLIS imaging over CSE techniques include its potential application in the MSK system at high fields, such as 7 T. Rather surprisingly, after more than two decades of development and hundreds of studies conducted at 7 T, only two MSK studies have been performed using the CSE technique (102,103). Both studies, published by the same MR group very recently, in 2024 and 2026, are methodological investigations that propose two different ways to overcome technical issues at 7 T. As a matter of fact, due to the very nature of CSE techniques, as the magnetic field increases there are exacerbations of CSE technical issues, including water-fat swapping and the need of very short ΔTEs necessary for water-fat separation. MUSLIS imaging -which actually borrows the features of the MP2RAGE, a sequence developed explicitly for high fields- might have its place in MSK imaging at 7 T. In general, the longer tissue T1 -characteristic of higher fields- represents an advantage for IR-based techniques; furthermore, the combination images might mitigate the signal inhomogeneities that characterize 7 T MR images. Overall, the CSE techniques -which are critically important in liver imaging with measurements of PDFF and T2*- are far to be established at high fields, both in MSK and liver; MUSLIS imaging has a potential to be an alternative to CSE for applications in MSK, especially at high fields such as 7 T.

Quantitative MRI, such as measurements of T1 relaxation times, was once limited to research studies but is now finding increasing applications in clinical practice and musculoskeletal imaging (104–114). Similarly to FLAWS imaging (53,54,115), which provides T1 maps of the brain, MUSLIS imaging yields T1 parametric maps of the MSK system. As matter of fact, the idea of the MUSLIS sequence stems from FLAWS imaging and both FLAWS and MUSLIS are based on the MP2RAGE, a sequence also designed to provide T1 maps. In future studies, MUSLIS imaging could be complemented by the acquisition of B1 maps to further improve the quality of the T1 parametric maps.

## Conclusion

The proof-of-concept of a novel approach to musculoskeletal imaging, MUSLIS MRI, is proposed here and demonstrated by numerical simulations and MRI experiments performed *in vivo* at 3 T. MUSLIS imaging provides i) lipid only images, ii) water only images, iii) combination images with signal properties (mitigation of signal inhomogeneities, for instance) that can be advantageous at high fields such as 7 T. In addition, MUSLIS imaging also yields T1 parametric maps of the musculoskeletal system.

## Data Availability

All data produced in the present study are available upon reasonable request to the authors.

## Supplementary Materials

*A subset of the 3D MUSLIS1 imaging volume.*

**Figure S1.**
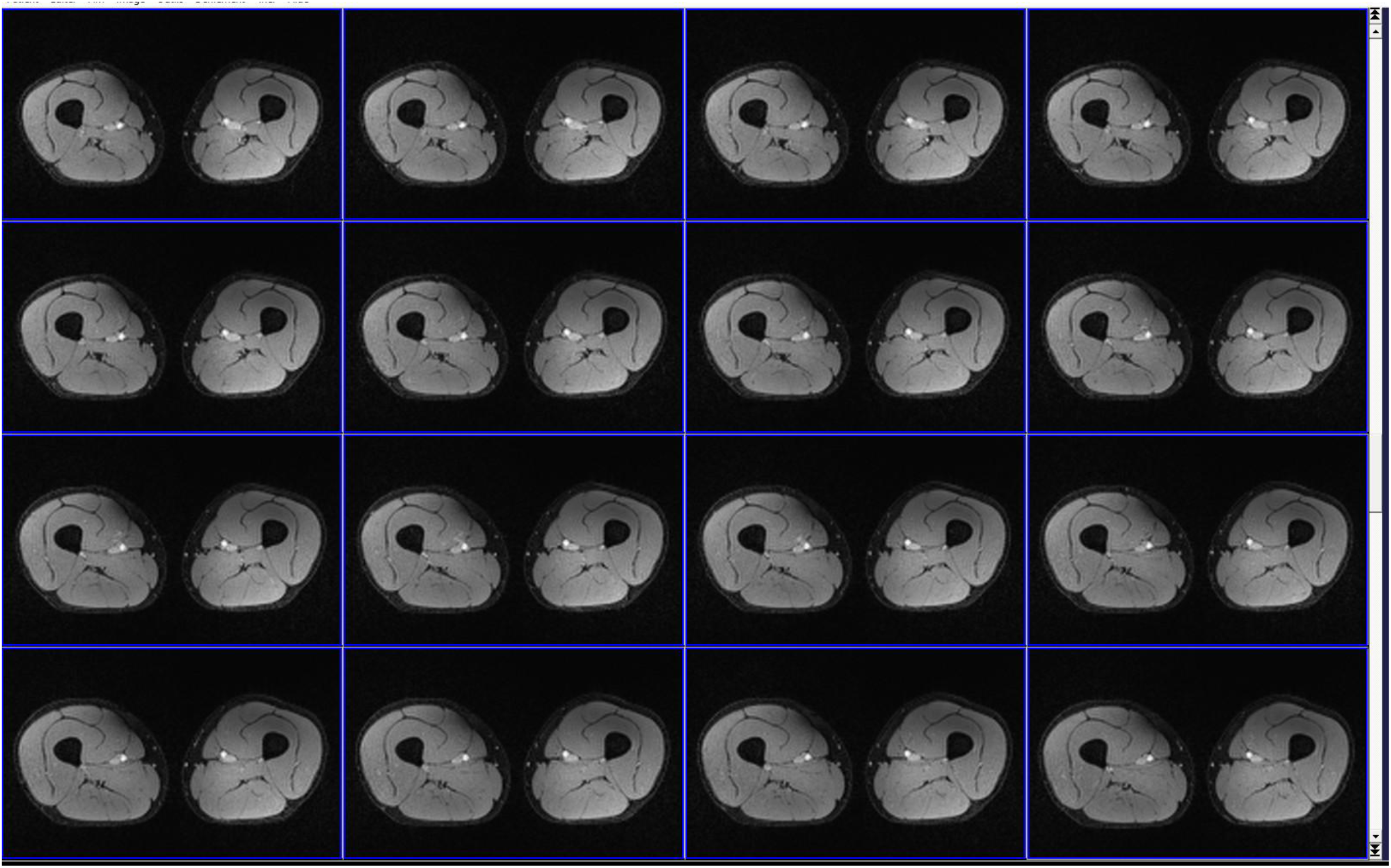
An example of a subset of slices from the MUSLIS1 imaging volume (sixteen contiguous slices out of the fifty-two slices). All slices of MUSLIS1 display a similar quality of lipid suppression.

*A subset of the 3D MUSLIS2 imaging volume.*

**Figure S2.**
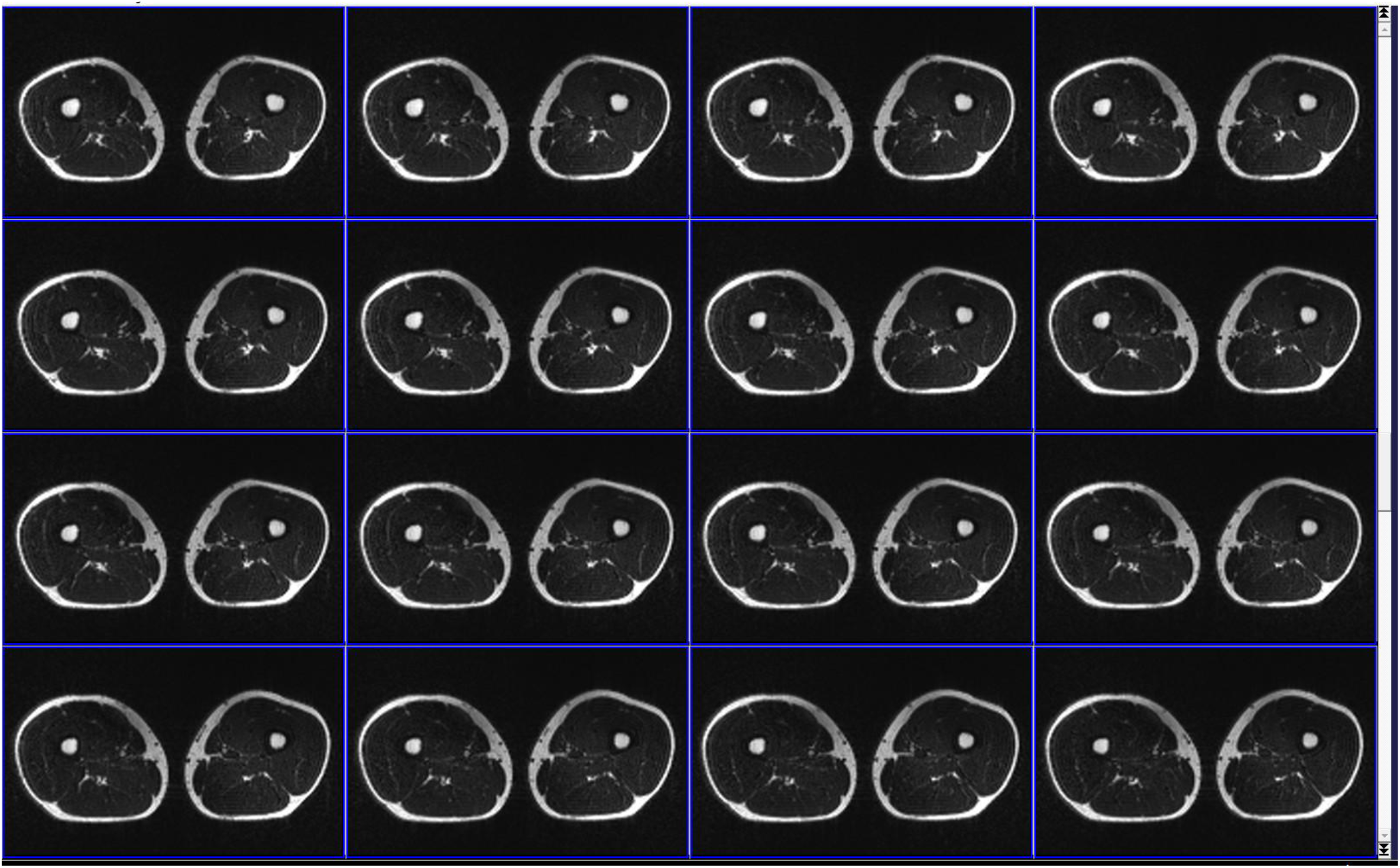
An example of a subset of slices from the MUSLIS2 imaging volume (sixteen contiguous slices out of the fifty-two slices). All slices of MUSLIS2 display a similar quality of muscle suppression.

*Subcutaneous adipose tissue: an example of signal intensity in the combination image MUSLIS-Max and in the acquired images MUSLIS1 and MUSLIS2.*

**Figure S3.**
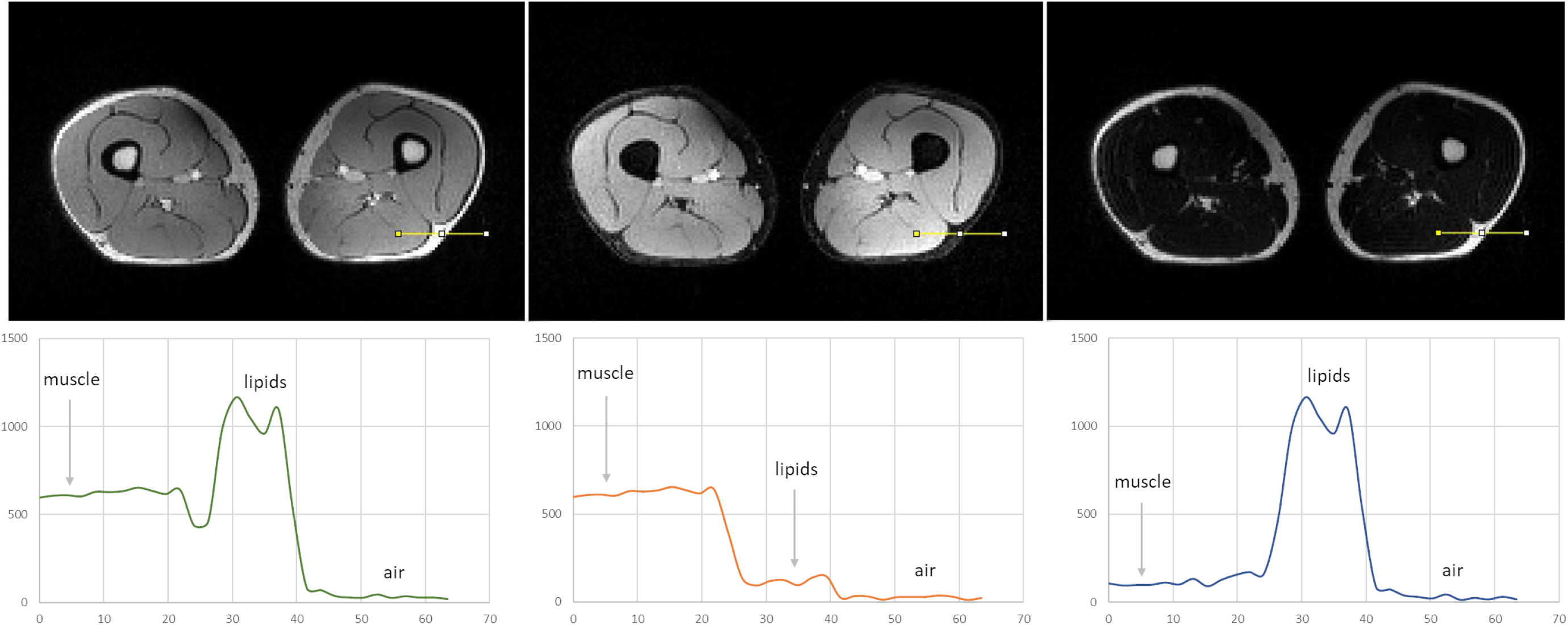
The lipid signal suppression and muscle signal suppression in MUSLIS imaging. **Upper panel**: the MUSLIS-Max (left), the MUSLIS1 (center) and the MUSLIS2 (right). For each image, the signal intensity is evaluated along the profile indicated by the yellow line. The profile is chosen to cover muscle, subcutaneous adipose tissue (lipids) and air. **Lower panel**: the signal intensity along the profile on MUSLIS-Max (left), MUSLIS1 (center) and MUSLIS2 (right). The value of signal intensity of lipids is close to the noise level (the signal in air) in MUSLIS1 (center). Similar observation applies to the signal intensity of muscle, which is close to the noise level, in MUSLIS2 (right). This figure well illustrates the excellent suppression of the target tissue (either lipid or muscle) with the intensity close to the value in air. This figure also illustrates the overall advantage of the max combination image. The same vertical scale is employed for the plots, for comparison purposes.

*Bone marrow adipose tissue: an example of signal intensity in the combination image MUSLIS-Max and in the acquired images MUSLIS1 and MUSLIS2.*

**Figure S4.**
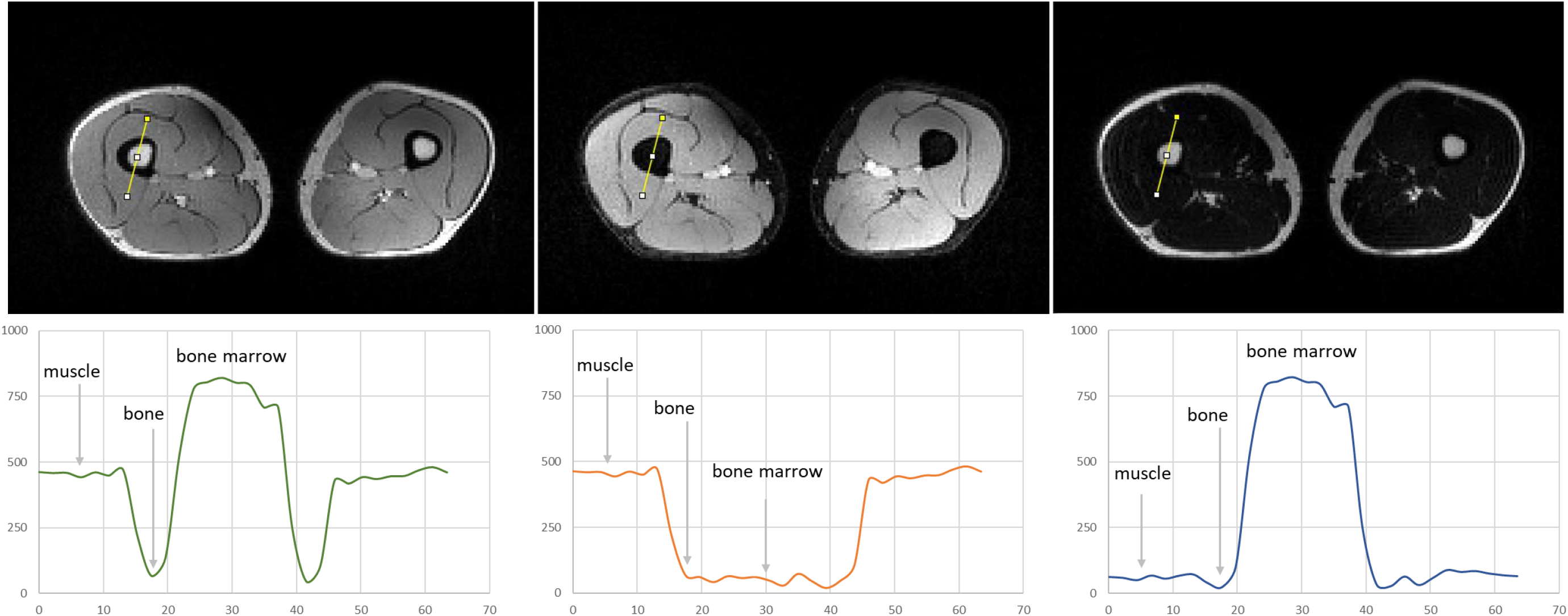
The lipid signal suppression and muscle signal suppression in MUSLIS imaging. MR images (**upper panel**:) and signal intensity along the a profile (**lower panel**) indicated by the yellow line. The profile is chosen to cover muscle, bone marrow adipose tissue (lipids) and cortical bone. This figure well illustrates the excellent suppression of the target tissue (either lipid or muscle) with the intensity close to the value in cortical bone. The same vertical scale is employed for the plots, for comparison purposes.

*Additional measurements of signal contrast in MUSLIS1.*

**Figure S5.**
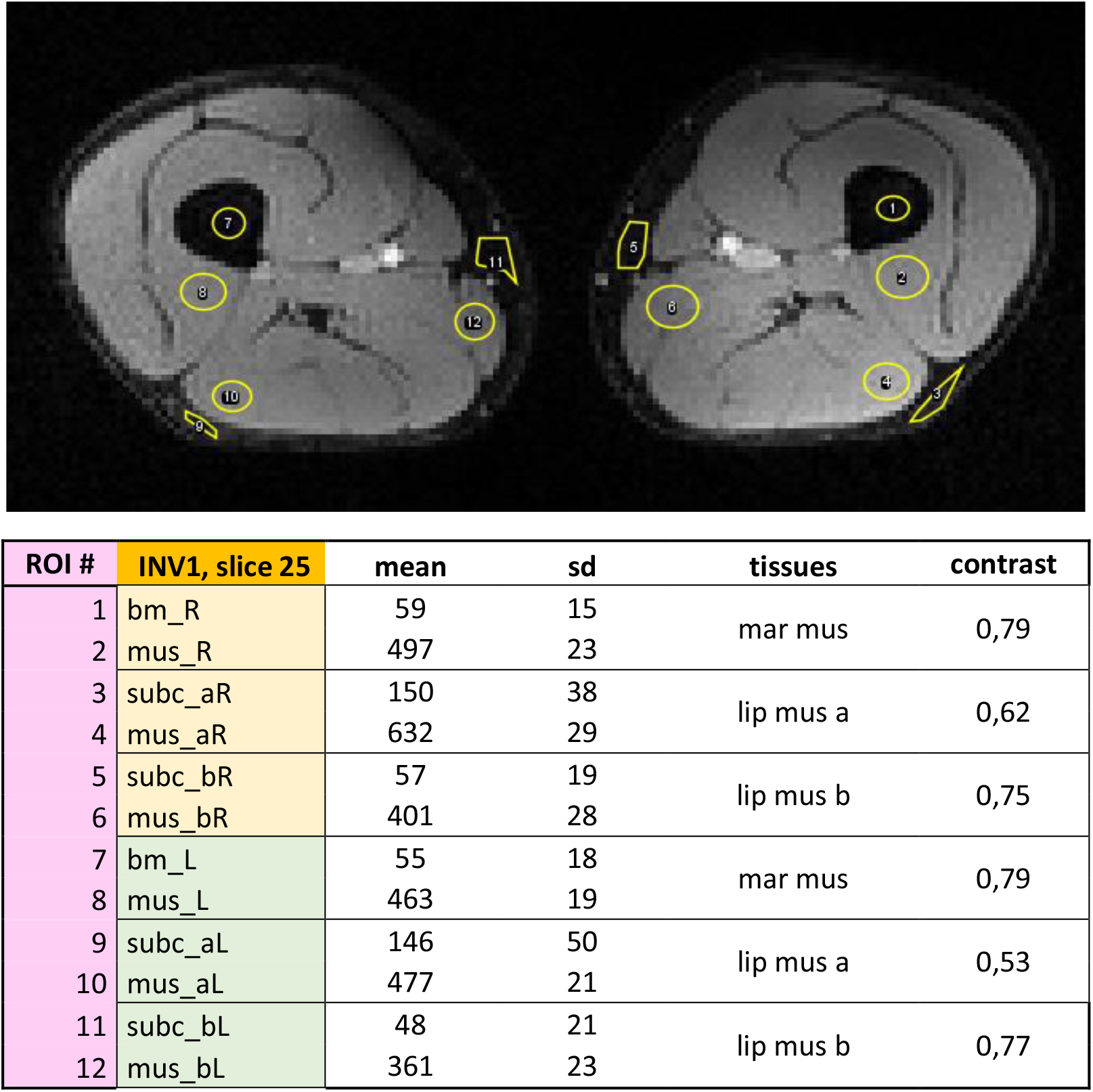
Example of MUSLIS1 image (**upper panel**). Table with the values of signal and contrast (**lower panel**) in the ROIs indicated on the image. Legend of abbreviations: bm_R = bone marrow, *right leg* (roi 1) mus_R = muscle, right leg (roi 2) subc_aR = subcutaneous fat, location a, *right leg* (roi 3) mus_aR = muscle, location a, *right leg* (roi 4) subc_bR = subcutaneous fat, location b, *right leg* (roi 5) mus_bR = muscle, location b, *right leg* (roi 6) bm_L = bone marrow, *left leg* (roi 7) mus_L = muscle, *left leg* (roi 8) subc_aL = subcutaneous fat, location a, *left leg* (roi 9) mus_aL = muscle, location a, *left leg* (roi 10) subc_bL = subcutaneous fat, location b, *left leg* (roi 11) mus_bL = muscle, location b, *left leg* (roi 12)

*Additional measurements of signal contrast in MUSLIS2.*

**Figure S6.**
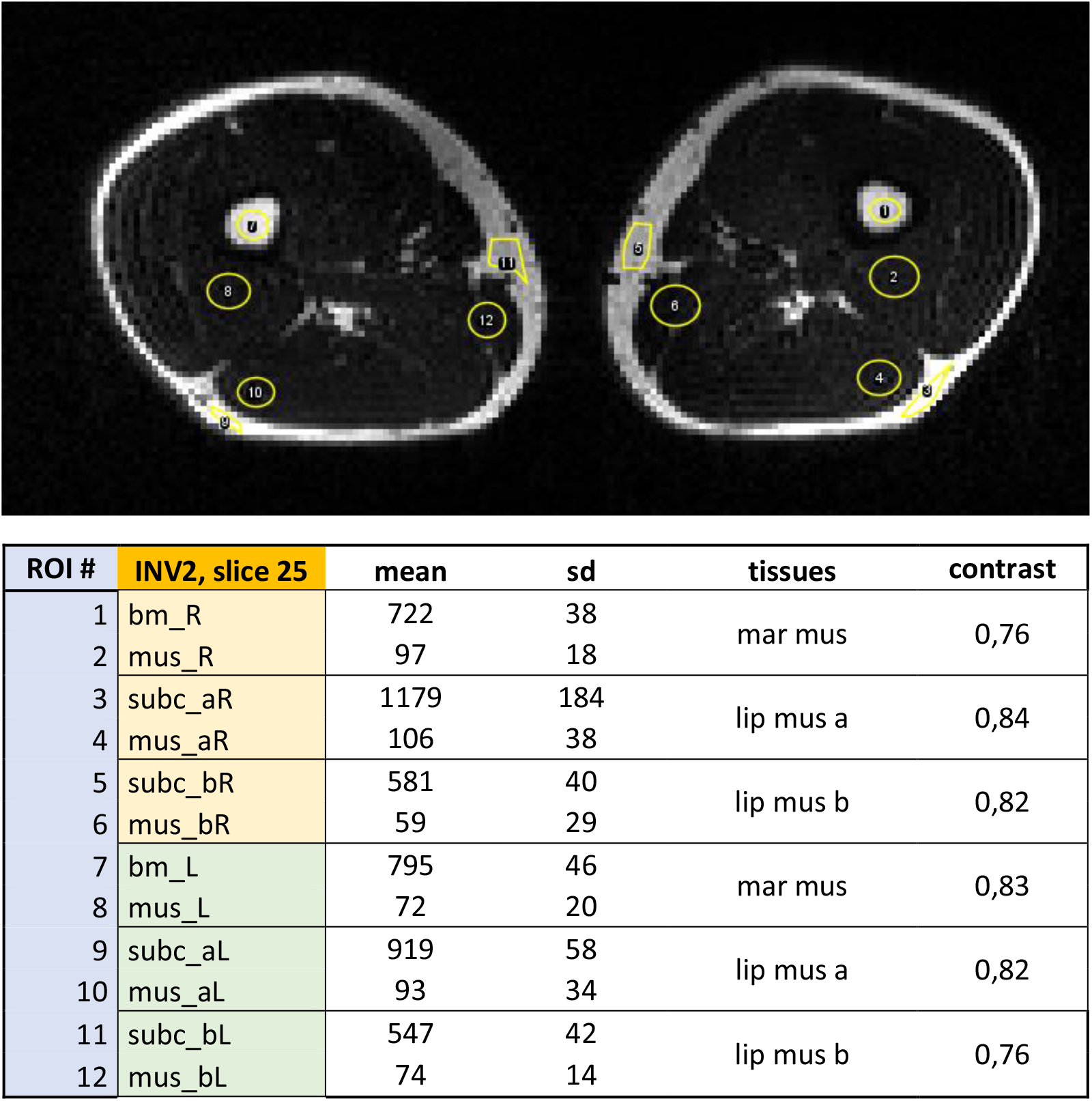
Example of MUSLIS2 image (**upper panel**). Table with the values of signal and contrast (**lower panel**) in the ROIs indicated on the image. Legend of abbreviations: bm_R = bone marrow, *right leg* (roi 1) mus_R = muscle, *right leg* (roi 2) subc_aR = subcutaneous fat, location a, *right leg* (roi 3) mus_aR = muscle, location a, *right leg* (roi 4) subc_bR = subcutaneous fat, location b, *right leg* (roi 5) mus_bR = muscle, location b, *right leg* (roi 6) bm_L = bone marrow, *left leg* (roi 7) mus_L = muscle, *left leg* (roi 8) subc_aL = subcutaneous fat, location a, *left leg* (roi 9) mus_aL = muscle, location a, *left leg* (roi 10) subc_bL = subcutaneous fat, location b, *left leg* (roi 11) mus_bL = muscle, location b, *left leg* (roi 12)

*Additional measurements of signal contrast in MUSLIS-Max.*

**Figure S7.**
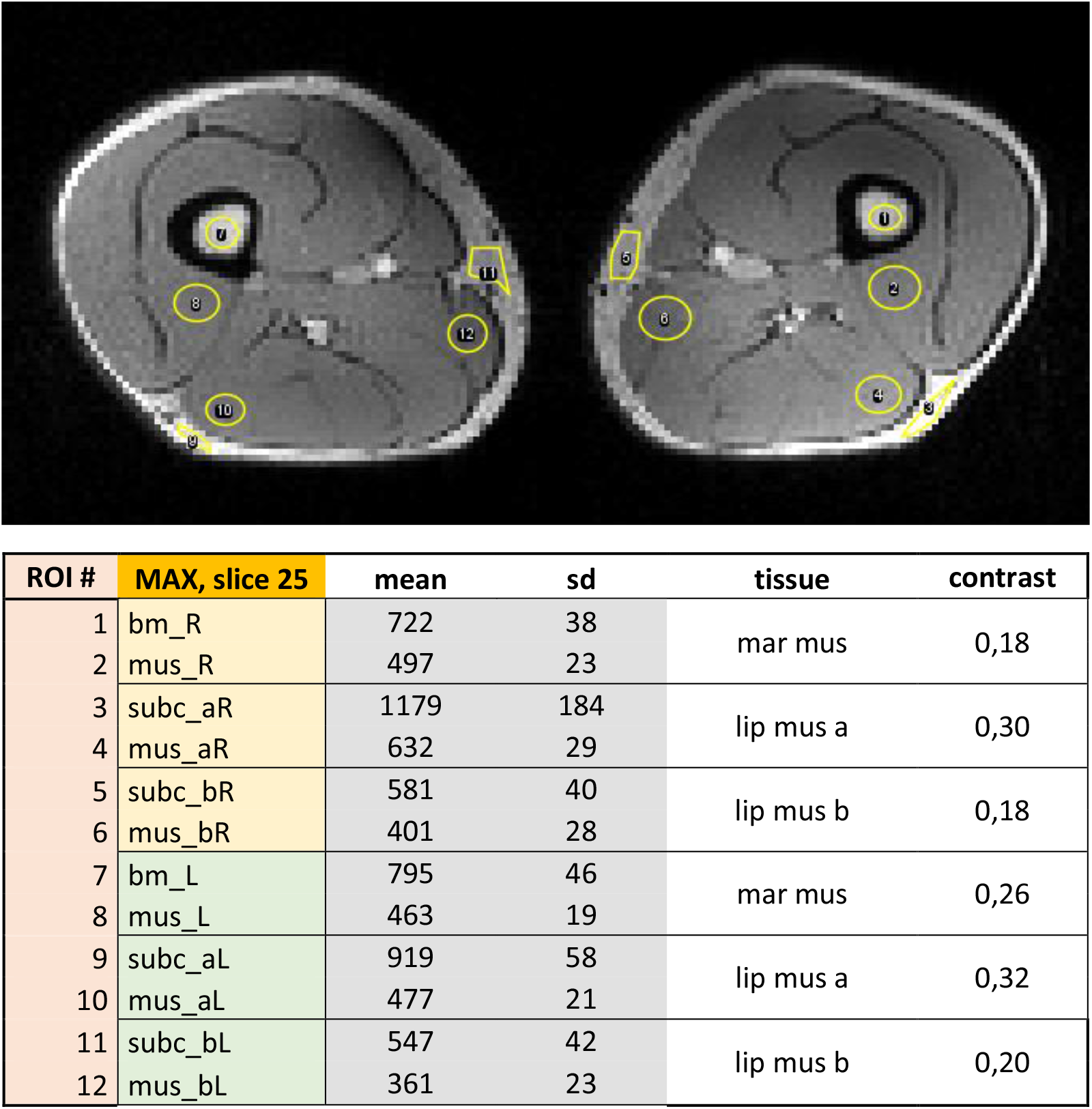
Example of MUSLIS-Max image (**upper panel**). Table with the values of signal and contrast (**lower panel**) in the ROIs indicated on the image. Legend of abbreviations: bm_R = bone marrow, *right leg* (roi 1) mus_R = muscle, *right leg* (roi 2) subc_aR = subcutaneous fat, location a, *right leg* (roi 3) mus_aR = muscle, location a, *right leg* (roi 4) subc_bR = subcutaneous fat, location b, *right leg* (roi 5) mus_bR = muscle, location b, *right leg* (roi 6) bm_L = bone marrow, *left leg* (roi 7) mus_L = muscle, *left leg* (roi 8) subc_aL = subcutaneous fat, location a, *left leg* (roi 9) mus_aL = muscle, location a, *left leg* (roi 10) subc_bL = subcutaneous fat, location b, *left leg* (roi 11) mus_bL = muscle, location b, *left leg* (roi 12)

